# Increased adiposity is protective for breast and prostate cancer: a Mendelian randomisation study using up to 132,413 breast cancer cases and 85,907 prostate cancer cases

**DOI:** 10.1101/2020.04.02.20049031

**Authors:** H. A. Amin, P. Kaewsri, A. M. Yiorkas, H. Cooke, A. I. Blakemore, F. Drenos

## Abstract

**Background:** Breast and prostate cancer are the first and second most common types of cancer in women and men, respectively. A recent campaign by Cancer Research UK emphasised obesity as being a causal risk factor for cancer, although previously published evidence is heterogenous. We aimed to explore the causal effect of adiposity on breast and prostate cancer risk in the UK Biobank (UKB), a large prospective cohort study, and published data.

**Methods:** We used Mendelian randomisation (MR) to assess the causal effect of body mass index (BMI), body fat percentage (BFP), waist circumference (WC), hip circumference (HC), and waist-to-hip ratio (WHR) on breast and prostate cancer risk.

**Results:** We obtained estimates (odds ratios, OR, per SD unit increase) of the causal effect of the adiposity measures on breast and prostate cancer risk. BMI and HC decrease the risk of breast cancer (OR 0.776 [95% CIs 0.661-0.91] and OR 0.781 [95% CIs 0.649-0.94], respectively). WC, BFP, and BMI decrease the risk of prostate cancer (OR 0.602 [95% CIs 0.439-0.825], OR 0.629 [95% CIs 0.414-0.956], and OR 0.695 [95% CIs 0.553-0.874], respectively). The protective effect of adiposity on prostate cancer risk is enhanced in men who are exposed to potentially hazardous substances at work, and the association between BMI and breast cancer is confounded by variables associated with general health.

**Conclusions:** In conclusion, increasing adiposity is causally protective for breast and prostate cancer and the effects in prostate cancer may, at least partly, be due to the safe storage of chemicals in adipose cells. It is necessary to explore the mechanisms through which adiposity may protect against or be a risk factor for cancer, to identify how the latter can be minimised without sacrificing the former, and to base public health campaigns around sound evidence.

**HIGHLIGHTS:**

- Previously published evidence regarding the effect of adiposity on prostate and breast cancer risk is heterogenous
- Increasing BMI and hip circumference decrease the risk of breast cancer in women
- Increasing waist circumference, body fat percentage, and BMI decrease the risk of prostate cancer
- The protective effect of adiposity on prostate cancer is stronger in men who are exposed to carcinogens at work
- Public health campaigns need to target the negative aspects of adiposity whilst preserving the positive aspects

## INTRODUCTION

Breast and prostate cancer are the most common and second most common types of cancer diagnosed worldwide in men and women, respectively.^1^ In 2010, the combined cost of breast and prostate cancer to the NHS was £664 million.^2^ The number of cases are expected to rapidly increase and, by 2040, are estimated to be 20.2% higher for breast cancer and 38.5% higher for prostate cancer in comparison to 2018.^1^ Both cancer types are preventable in many cases, making robust identification of their modifiable risk factors important.

A recent campaign by Cancer Research UK^3^ has emphasised obesity as being a causal risk factor for cancer comparable to smoking. It has been proposed that the metabolic environment in obese people is conducive to oncogenic transformation.^4^ However, previously published evidence on the relationship between adiposity and breast and prostate cancer does not consistently support this view.^5-7^ It has been suggested that adiposity is a risk factor for breast cancer in post-menopausal women,^8^ but not in pre-menopausal women.^9^ The effect of BMI on prostate cancer risk is reported to depend upon the aggressiveness of the tumour.^10^

Assessment of what exposures are causal is not trivial: “correlation is not causation”. The vast majority of studies carried out to examine the impact of adiposity on breast and prostate cancer risk are observational and may be susceptible to confounding. Mendelian randomisation (MR) is a method that uses genetic variants associated with an exposure of interest, but not with any confounders, to assess the causal effect of the genetically predicted exposure on an outcome. In order for the method to provide reliable estimates of the causal effect, it is also assumed that the chosen instruments are not related to the outcome of interest independently of the exposure (this is known as the “exclusion restriction” assumption).

With this work, we aim to explore the causal effect of adiposity on breast and prostate cancer risk in the UK Biobank (UKB), a large prospective cohort study, and published data.^11-15^ We also aim to use the rich phenotype data collected as part of the UKB study to identify variables, if any, that may explain the observed relationship between adiposity and breast and prostate cancer risk.

## SUBJECTS AND METHODS

### Population and Study Design

The UK Biobank (UKB) is a large prospective cohort study including information and biological samples for approximately 500,000 individuals, recruited between 2006 and 2011. The 22 UKB assessment centres throughout England, Wales and Scotland, collected baseline data from the participants in the form of questionnaires, physical and cognitive tests and blood and urine samples.^16^ The age range of the participants at the time of enrolment in the study was between 40 and 69 years of age, with a mean age of 56.5 years. Males represent 45.6% of the sample. The use of the data for this project was approved by the UKB (application 44566).

### Genotyping

488,377 individuals had been genotyped for up to 812,428 variants using DNA extracted from blood samples on either the UKB Axiom array (438,427 participants) or the UK BiLEVE Axiom array (49,950 participants). Variants that did not pass standard quality control checks were excluded.^17^ These included tests for the presence batch effects, plate effects, sex effects and array effects, as well as any departures from Hardy-Weinberg Equilibrium using a p-value threshold of 10^−12^. Variants with a minor allele frequency of <0.01 were also excluded. For imputed variants, all variants with an INFO score of <0.8 were excluded from the analysis.

Sample genotyping quality control metrics were provided by UKB.^17^ Samples were excluded from the analysis if they were outliers for missingness and/or PC-corrected heterozygosity and/or if they had any sex chromosome aneuploidies as well as if the genetically inferred sex differed from the reported sex. Samples which did not have a genetically determined White British ancestry were also excluded. A list of related individuals was also provided by UKB and one individual from each related pair was excluded at random.

### Phenotypes

We used data collected at baseline for body mass index (BMI, UKB field 21001), body fat percentage (BFP, UKB field 23099) from bio-impedance, waist circumference (WC, UKB field 48) and hip circumference (HC, UKB field 49). We calculated waist-to-hip ratio (WHR) by dividing WC by HC. The variables were standardised to a mean of 0 and a variance of 1.

We used cancer diagnoses information from the 1970s onward obtained from linkage to national cancer registries and health records (see http://biobank.ndph.ox.ac.uk/showcase/showcase/docs/CancerLinkage.pdf). Breast cancer cases are defined as females who have an ICD-10 code C50 recorded at least once (UKB field 40006). Prostate cancer cases are defined as males who have an ICD-10 code C61 recorded at least once (UKB field 40006). Females who have an ICD-10 code D05, for in situ carcinoma of the breast, without a C50 breast cancer entry were removed from the sample. Similarly, males with an ICD-10 code D075, for carcinoma in situ of prostate, without a C61 prostate cancer diagnosis were also removed from the sample.

Menopause information for females was obtained through the reported age of menopause information collected (UKB field 3581). This information was compared to the age of first breast cancer diagnosis to identify the pre- and post-menopausal cases. For women who did not have breast cancer, we used their menopause status at baseline to stratify them into pre- and post-menopausal.

Exposure to chemicals was based on occupation exposure information. A participant was considered to have been exposed to chemicals frequently if they answered “Often” and/or “Sometimes” at least once for any of the following UKB fields: 22609 (Workplace very dusty); 22610 (Workplace full of chemical or other fumes); 22611 (Workplace had a lot of cigarette smoke from other people smoking); 22612 (Worked with materials containing asbestos); 22613 (Worked with paints, thinners or glues); 22614 (Worked with pesticides); and 22615 (Workplace had a lot of diesel exhaust).

### Statistical Analyses

We used R 3.6.1^18^ to carry out analyses and generate plots, unless stated otherwise. For the observational analyses, we removed prevalent cases and regressed the exposures (i.e. BMI, BFP, WC, HC, and WHR) against prostate and breast cancer cases using a logistic regression adjusting for age at baseline. We then generated genetic risk scores (GRS) for the exposures in PLINK 1.9^19^ using sex-specific summary statistics from GIANT^14, 15^ and Lu *et al*.^11^ We tested the association between the GRSs and prostate and breast cancer adjusting for the first four principal components (PCs) for the genetic variability of the genome, age at baseline, and genotyping array used. We stratified the sample by menopause and by chemical exposure and repeated this analysis. We used the GRSs as instruments to carry out MR as described elsewhere.^20^ We adjusted for the first four genetic PCs and the genotyping array used in the first step and adjusted for age at baseline in the second step.

Two-sample MR and multivariate MR analyses were carried out using the TwoSampleMR R package.^21^ All of the independent loci used as instruments in the two-sample MR analyses were based on sex-stratified GWAS meta-analyses of European ancestry. For BFP, we used the sex-stratified summary statistics for 10 variants obtained from Lu *et al*.^11^ We obtained sex-stratified summary statistics from GIANT for waist-to-hip ratio, waist circumference, hip circumference,^14^ and BMI.^15^ If a variant did not reach genome-wide statistical significance for men or women in the sex-stratified analyses in the external data, it was excluded. We used the same information for the multivariate two-sample MR with the exception of BFP sex-stratified summary statistics, which were not available, and the estimates were generated using UKB data instead. The outcome data used were from Michailidou *et al*.^12^ and Shumacher *et al*.,^13^ which are the meta-analysis of breast cancer (122,977 cases, 105,974 controls) and the meta-analysis of prostate cancer (79,194 cases, 61,112 controls), respectively.

To identify the potential confounders, we developed an algorithm that used a step-wise procedure to identify any variables in the UKB that may be responsible for the observed positive association between BMI and incident breast cancer cases. Only variables with more than 1000 non-missing observations associated with both BMI and breast cancer (p<0.05) were considered. Categorical phenotypes were converted to separate binary variables. Our algorithm selected variables based on the extent to which they minimised the effect size of BMI on breast cancer.

## RESULTS

After QC, we had 151,940 males and 174,410 females remaining in the sample. Table 1 summarises their age, BMI at baseline, and lifetime smoking status.

**Table 1.** Population characteristics. SD = standard deviation.

|  | **males** |  |  | **females** |  |  |
| --- | --- | --- | --- | --- | --- | --- |
|  | **all** | **controls** | **cases** | **all** | **controls** | **cases** |
| n | 151940 | 145227 | 6713 | 174410 | 164974 | 9436 |
| age (SD), years | 57.1 (8.08) | 56.8 (8.1) | 63 (4.88) | 56.6 (7.9) | 56.5 (7.92) | 58.9 (7.1) |
| BMI (SD), kg/m<sup>2</sup> | 27.8 (4.22) | 27.8 (4.24) | 27.6 (3.83) | 27 (5.14) | 27 (5.15) | 27.3 (4.93) |
| % ever smoked | 65.1 | 65 | 66.1 | 55.7 | 55.6 | 57.8 |

We first sought to examine the observational effect of the adiposity measures (i.e. BMI, BFP, WC, HC, and WHR) on the risk of breast and prostate cancer. We only used incident cases to avoid a previous cancer diagnosis affecting any of the measures considered. We found that, in our sample, each of the adiposity measures are associated with an increased risk of breast cancer but a decreased risk of prostate cancer (Figure 1 & Supplementary Table 1).

**Figure 1.**
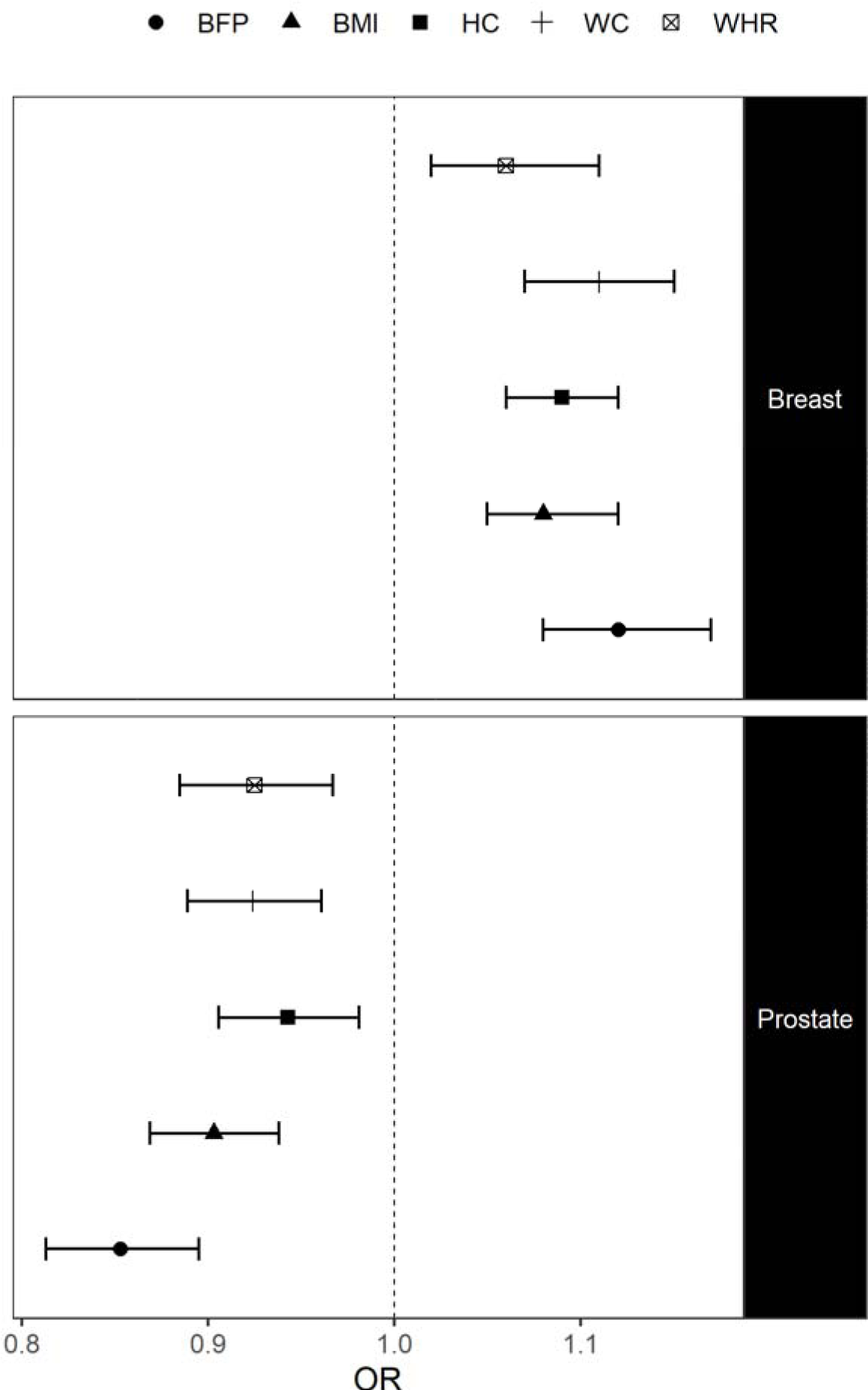
Odds ratios per SD increase (OR) and 95% confidence intervals when regressing incident breast and prostate cancer cases on the adiposity measures using a logistic regression. BMI = body mass index. BFP = body fat percentage. HC = hip circumference. WC = waist circumference. WHR = waist-to-hip ratio.

We generated GRSs for each of the adiposity measures, for males and females separately, to use as instrumental variables (Supplementary Table 2). We regressed the outcomes on each of these GRSs to determine whether males and females who are genetically predisposed to increased adiposity are at a decreased or increased risk of developing prostate or breast cancer, respectively. We found that, in our sample, a genetic predisposition to increased BMI, WC, and HC is associated with a decreased risk of breast cancer in women, and that a genetic predisposition to increased BMI and WC is associated with a decreased risk of prostate cancer (Supplementary Figures 1-3 & Supplementary Table 3).

We found that the associations between the GRSs for BMI, WC, and HC and breast cancer were present even after adjusting for BMI, WC, and HC, respectively (Supplementary Table 4). This suggests that these GRSs violate the exclusion restriction assumption, i.e. the genetic instrument may affect the outcome independently of the exposure, end the estimated causal effect may be biased. We addressed this issue by confirming our one-sample MR results using the MR Egger method, which is robust even when the exclusion restriction assumption is violated. We found that the GRS for BMI was not associated with prostate cancer independently of BMI. However, the GRS for WC was associated with prostate risk independently of WC (Supplementary Table 4), so the estimate of the causal effect of WC on prostate cancer risk obtained in the UKB may also be biased.

We obtained estimates (odds ratios, OR, per SD unit increase) of the causal effect of the adiposity measures on breast and prostate cancer using one-sample MR (Figures 2 & 3). We found that increased BMI and HC decreased the risk of breast cancer (OR 0.776 [95% CIs 0.661 to 0.91, p = 1.79×10^−3^] and OR 0.781 [95% CIs 0.649 to 0.94, p = 9.02×10^−3^], respectively), and increased WC, BFP, and BMI decreased the risk of prostate cancer (OR 0.602 [95% CIs 0.439 to 0.825, p = 1.61×10^−3^], OR 0.629 [95% CIs 0.414 to 0.956, p = 0.0301], and OR 0.695 [95% CIs 0.553 to 0.874, p = 1.89×10^−3^], respectively).

**Figure 2.**
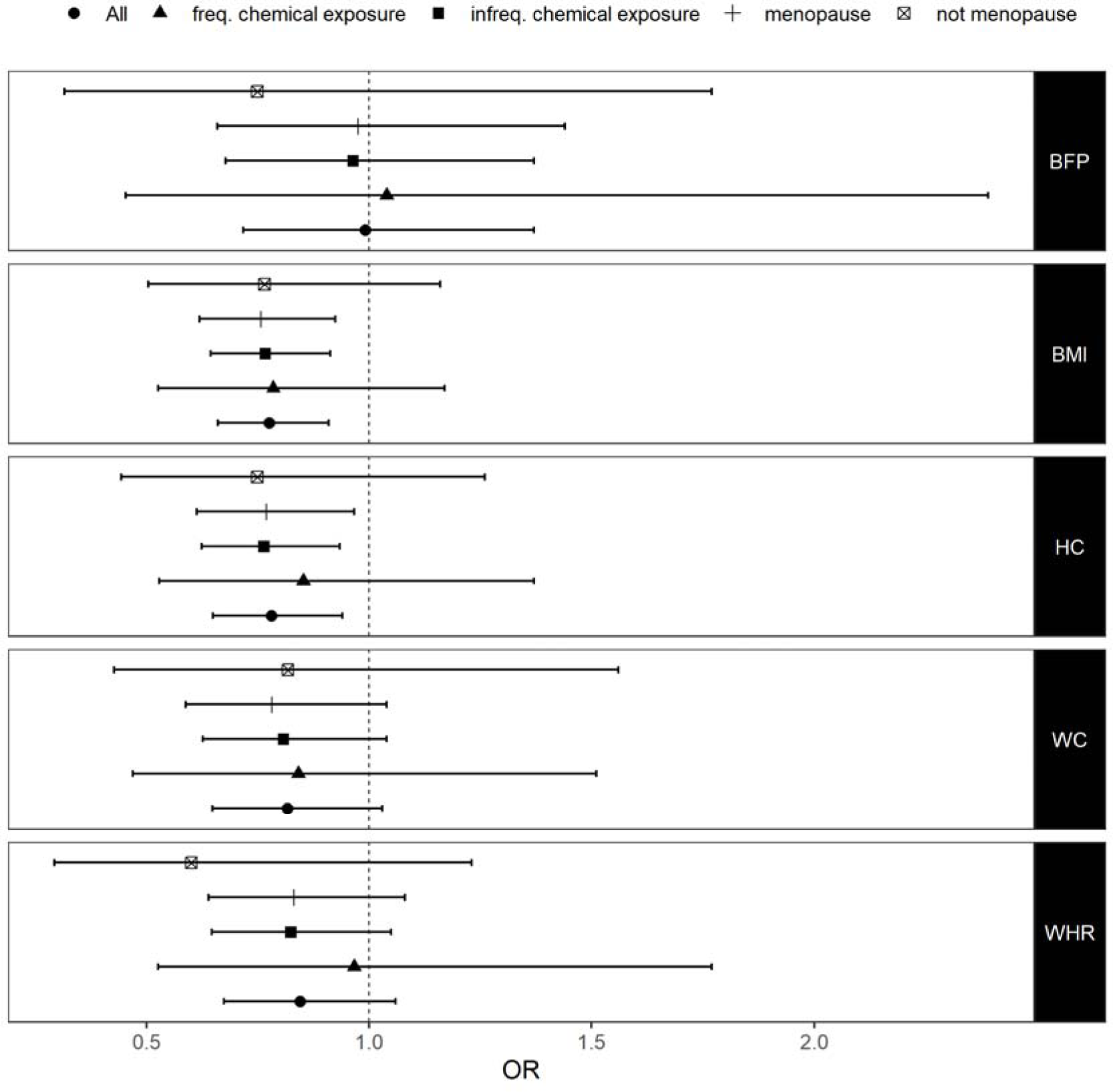
Odds ratios (OR) and 95% confidence intervals from a one-sample MR analysis of the causal effect of adiposity on breast cancer risk. BMI = body mass index. BFP = body fat percentage. HC = hip circumference. WC = waist circumference. WHR = waist-to-hip ratio.

We sought to replicate our findings using external outcome summary statistics from meta-analyses of 122,977 breast cancer cases^12^ and 79,194 prostate cancer cases,^13^ and external exposure summary statistics from Lu *et al*.^11^ and GIANT.^14, 15^ We used two-sample MR to assess the causal effect of adiposity measures on breast and prostate cancer risk (Tables 2 & 3). We found that increased BMI, WC, HC, and BFP are causally protective for breast cancer using the inverse variance weighted method (Supplementary Figure 4). The p-values of the intercept from the MR Egger method suggest that the instruments used for BMI, WC, and HC may be pleiotropic, but the causal estimates generated using MR Egger show that increased BMI, WC, and HC are still protective for breast cancer when any bias from pleiotropy is taken into account (Table 2). We also found that BMI was the only adiposity measure causally protective for prostate cancer (Supplementary Figure 5).

**Table 2.** Odds ratios (OR) and 95% confidence intervals from a two-sample MR analysis of the causal effect of adiposity on breast cancer risk in women. U 95% CI = upper 95% confidence interval. L 95% CI = lower 95% confidence interval. *P*^(intercept)^ = p-value of the intercept from the MR Egger method. BMI = body mass index. BFP = body fat percentage. HC = hip circumference. WC = waist circumference. WHR = waist-to-hip ratio.

| Cancer | Exposure | Method | *P* | OR | L 95% CI | U 95% CI | $P^{(\text{intercept})}$ |
| --- | --- | --- | --- | --- | --- | --- | --- |
| Breast | BMI | MR Egger | 2.32E-05 | 0.444 | 0.321 | 0.616 | 0.00839 |
| Breast | BMI | Inverse variance weighted | 3.63E-08 | 0.683 | 0.596 | 0.782 | NA |
| Breast | BFP | MR Egger | 0.221 | 0.467 | 0.152 | 1.44 | 0.59 |
| Breast | BFP | Inverse variance weighted | 0.0215 | 0.63 | 0.425 | 0.934 | NA |
| Breast | HC | MR Egger | 0.00414 | 0.276 | 0.129 | 0.587 | 0.0236 |
| Breast | HC | Inverse variance weighted | 0.00725 | 0.689 | 0.525 | 0.904 | NA |
| Breast | WC | MR Egger | 3.64E-05 | 0.291 | 0.192 | 0.443 | 0.00124 |
| Breast | WC | Inverse variance weighted | 2.71E-05 | 0.643 | 0.523 | 0.79 | NA |
| Breast | WHR | MR Egger | 0.631 | 0.7 | 0.168 | 2.91 | 0.747 |
| Breast | WHR | Inverse variance weighted | 0.112 | 0.807 | 0.62 | 1.05 | NA |

**Table 3.** Odds ratios (OR) and 95% confidence intervals from a two-sample MR analysis of the causal effect of adiposity on prostate cancer risk. U 95% CI = upper 95% confidence interval. L 95% CI = lower 95% confidence interval. *P*^(intercept)^ = p-value of the intercept from the MR Egger method. BMI = body mass index. BFP = body fat percentage. HC = hip circumference. WC = waist circumference. WHR = waist-to-hip ratio.

| Cancer | Exposure | Method | OR | L 95% CI | U 95% CI | P | $P^{(\text{intercept})}$ |
| --- | --- | --- | --- | --- | --- | --- | --- |
| Prostate | BMI | MR Egger | 0.841 | 0.623 | 1.13 | 0.266 | 0.828 |
| Prostate | BMI | Inverse variance weighted | 0.815 | 0.73 | 0.91 | 0.000278 | NA |
| Prostate | BFP | MR Egger | 1.01 | 0.291 | 3.53 | 0.985 | 0.882 |
| Prostate | BFP | Inverse variance weighted | 0.921 | 0.696 | 1.22 | 0.563 | NA |
| Prostate | HC | MR Egger | 0.703 | 0.248 | 2 | 0.522 | 0.706 |
| Prostate | HC | Inverse variance weighted | 0.858 | 0.661 | 1.11 | 0.252 | NA |
| Prostate | WC | MR Egger | 1.39 | 0.203 | 9.53 | 0.745 | 0.664 |
| Prostate | WC | Inverse variance weighted | 0.801 | 0.604 | 1.06 | 0.123 | NA |
| Prostate | WHR | MR Egger | 0.482 | 0.183 | 1.27 | 0.277 | 0.23 |
| Prostate | WHR | Inverse variance weighted | 0.842 | 0.659 | 1.08 | 0.17 | NA |

We next performed multivariable MR to identify whether the protective effects of increased BMI, WC, HC, and BFP on breast cancer risk are independent of each other (Supplementary Table 6). We found that BMI and BFP were still protective for breast cancer independently of the other measures, but WC and HC were not.

We hypothesised that the protective effect of adiposity may be due to adipose tissue absorbing and safely storing environmental carcinogens. We, therefore, stratified our sample, based on self-reported exposure to dust and/or chemicals and/or fumes at work and repeated the analyses. We found that the protective effect of increasing adiposity on prostate cancer was stronger in men who reported that they were frequently exposed to potentially hazardous substances at work in comparison those who were not. This is particularly evident for BMI and WC, where the confidence intervals of the frequently and infrequently exposed strata do not overlap (Figure 3). We did not observe the same pattern in women (Figure 2).

**Figure 3.**
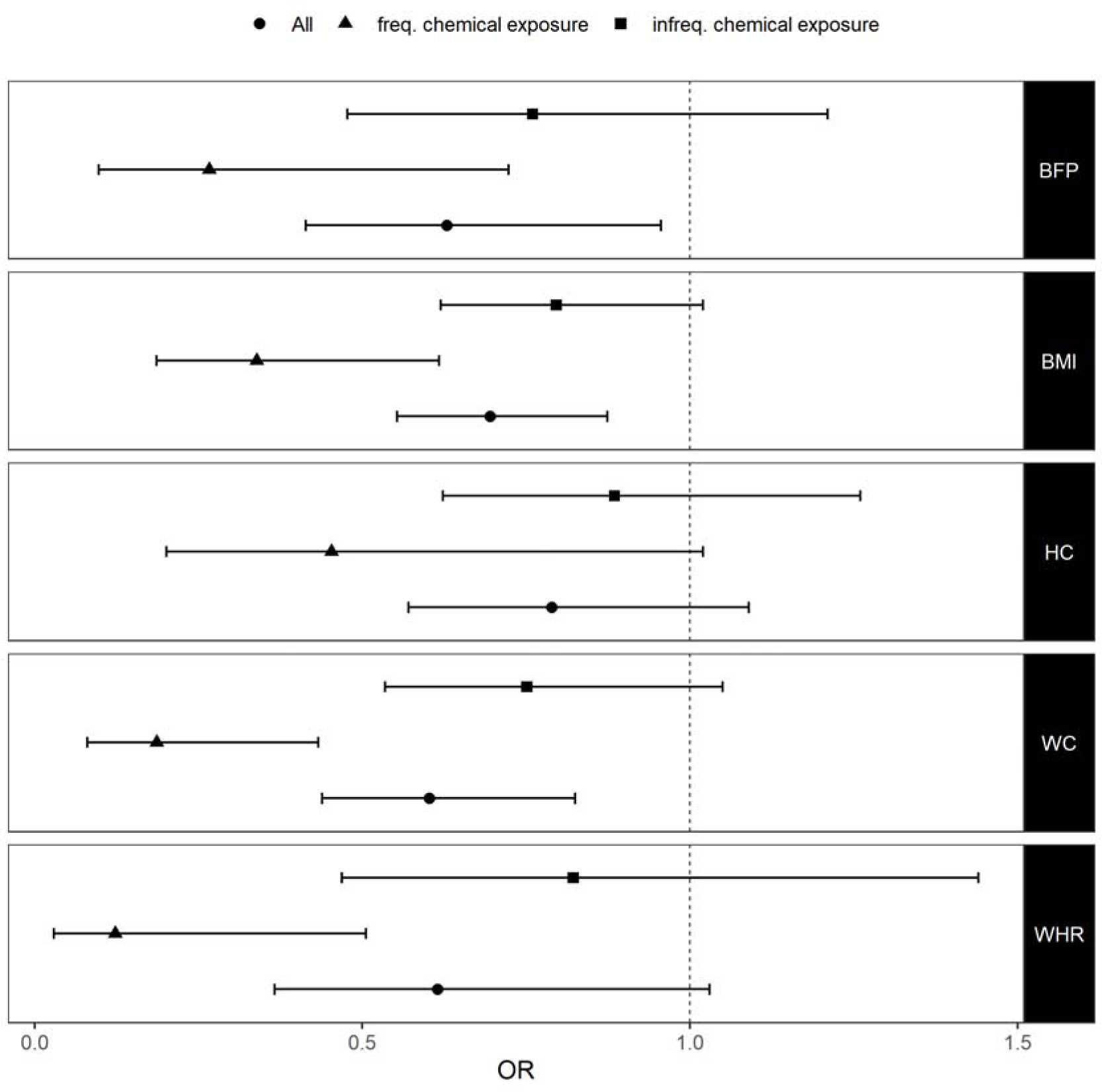
Odds ratios (OR) and 95% confidence intervals from a one-sample MR analysis of the causal effect of adiposity on prostate cancer risk. BMI = body mass index. BFP = body fat percentage. HC = hip circumference. WC = waist circumference. WHR = waist-to-hip ratio.

We stratified women based on their menopause status at breast cancer diagnosis (or at baseline for controls). We did not find evidence that the protective effect of adiposity was different between pre- and post-menopausal women, with the estimates between the groups being similar and having overlapping confidence intervals (Figure 2).

We used a stepwise procedure to identify any confounding variables that might explain the opposite direction of effects estimated by the observational and MR associations of BMI with risk of breast cancer. Supplementary Table 7 lists the fields that were both associated with incident breast cancer risk and attenuated the detrimental effect of BMI. The ln(OR) of breast cancer per SD unit increase in BMI is reduced tenfold when these variables (namely ankle spacing width, frequency of stair climbing, amount of moderate physical activity, macular degeneration, and leukocyte count) are added to the model, but it does not decrease below zero. It is possible that the variables our algorithm selects may be associated with a missing or currently unknown higher order variable that may explain the discrepancy between the observed and causal associations between BMI and risk of breast cancer.

## DISCUSSION

We sought to assess the causal effects of increased adiposity on the risk of breast and prostate cancer. We found that increased adiposity measures were observationally associated with a lower risk of prostate cancer, but with an increased risk of female breast cancer. When we assessed the causal effects of the adiposity measures on the risk of prostate and breast cancer using a one sample MR, we found that increased BMI, BFP, and WC were causally protective for prostate cancer, and increased BMI and HC were causally protective for breast cancer. Using a two sample MR with previously published data, BMI was further confirmed as causally protective for both outcomes, and HC was confirmed as causally protective for breast cancer. Multivariable MR analyses suggest that BFP and BMI are the independent drivers of these protective associations between the adiposity measures and breast cancer. When testing whether or not these protective effects could be attributed to the safe storage of chemicals in adipose tissue, we showed that the causally protective effect on prostate cancer was consistently stronger, though not always statistically significant, in those reporting exposure to potentially carcinogenic substances at work. However, no such association was evident for breast cancer. When we attempted to identify the confounders responsible for the observed detrimental association between increasing BMI and breast cancer, we found a number of variables that may be involved, but these are of a currently uncertain clinical significance.

In this work, we found that increases in all of the adiposity measures we tested were observationally associated with a higher number of breast cancer cases. In this respect, the UK Biobank is in agreement with a previously published large meta-analysis of 126 studies finding the same association.^22^ The inverse associations between the adiposity measures and prostate cancer were more surprising. Here the evidence are more heterogeneous, as illustrated by a recent large scale meta-analysis,^10^ which found an overall null association between BMI and prostate cancer, but found an inverse association between BMI and prostate specific antigen concentrations. Furthermore, a number of well powered studies^23, 24^ have also identified an inverse association between BMI and prostate cancer, so our results in the UK Biobank are, therefore, not unusual. Furthermore, increased adiposity is only protective for low grade prostate cancer,^10^ the prostate cancer cases in the UKB are likely to be low grade due to the age of the sample, and this may further explain why we found an inverse observational relationship between adiposity and prostate cancer risk.

Since observational studies cannot directly provide information on cause and effect relationships, we carried out MR analyses to see whether the associations we found were causal. We found that adiposity was causally protective for breast cancer and our results are similar to those reported by Guo *et al*.^6^ using the same study but with a lower number of cases. Observationally, adiposity has been reported to be protective for pre-menopausal breast cancer,^9^ but our results suggest that this protective effect can only be established for post-menopausal breast cancer cases, though the absence of a protective effect in pre-menopausal breast cancer cases may be due to the smaller number of cases in this cohort. We show that the causal estimate of the protective effect of body fat percentage on breast cancer is independent of the causal effect of BMI, and it is more reliable because we find no evidence that the GRS for BFP is pleiotropic. We also found evidence that the association between BMI and breast cancer is confounded, and that the confounder is likely to be either a non-observed variable or a higher order variable combining different existing variables in the data. We also found that BMI is causally protective for prostate cancer in the UK Biobank dataset, and this is supported by our analysis of external data. Davies *et al*.^7^ report no causal effect of BMI on prostate cancer, but this may be due to lack of statistical power because they used a smaller number of cases (20,848 vs. 79,194).

There is previously published evidence which suggests that adipose tissue may play a role in safely storing harmful chemicals.^25^ Persistent organic pollutant (POP) concentrations increase by 2-4% per kg of weight loss and remain elevated for up to 12 months after a weight loss intervention.^26^ We hypothesised that the protective effect of increasing adiposity on prostate and breast cancer risk might be explained by its ability to sequester potentially carcinogenic substances. Our results, which show that the protective effect was enhanced in men reporting more frequent exposure to potentially carcinogenic substances at work, support our hypothesis in prostate cancer. The same effect was not observed in female breast cancer, which may be due to an insufficient number of cases or due to a more complex underlying mechanism.

The direction of the observational association between BMI and breast cancer is opposite to that of the causal effect, which suggests that the former is confounded. We found that variables relating to physical activity (i.e. frequency of stair climbing and moderate physical activity) may be one source of confounding and this is supported by the fact that increased physical activity is protective for breast cancer^27^. Our algorithm also selected macular degeneration (an eye disease for which increasing age is the strongest risk factor and circulating lipids have also been involved),^28^ ankle width (which might represent swelling of the lower extremities – symptoms of diabetes and cardiovascular disease), and leukocyte count (a marker of systemic inflammation).^29^ These variables are likely to represent a currently undefined higher order variable, perhaps biological age or a marker of overall health, and further investigation is required to identify what this variable might be and whether or not it can be modified to minimise breast cancer risk.

A number of limitations are present in our work. The UK Biobank study, despite its sample size and almost comprehensive phenotyping, does have a “healthy volunteer” selection bias. The rate of cancer is lower in comparison to the general population.^30^ Also, the proportion of adults who were overweight or obese among men and women in the UK population was 78% and 73%, respectively, compared to 74% and 60%, respectively, for the same age group in the UK Biobank.^31^ The sample is, therefore, not representative of adiposity in the wider UK population.

In conclusion, we found that increased adiposity is causally protective for breast and prostate cancer and the effects in the prostate cancer, at least partly, may be due to the safe sequestration of chemicals in adipose cells. Further work needs to be done to identify variables that are responsible for the observed relationship between increased BMI and increased risk of breast cancer. It is clear that reduction of adiposity, in and of itself, may not reduce the risk of breast and prostate cancer as the recent campaign by Cancer Research UK^3^ might suggest. It is necessary to explore the mechanisms through which adiposity may protect against or be a risk factor for cancer, to identify how the latter can be minimised without sacrificing the former, and to base public health campaigns around sound evidence.

## Data Availability

This research has been conducted using the UK Biobank Resource under project 44566 (https://www.ukbiobank.ac.uk/2018/12/genetic-and-non-genetic-factors-able-to-predict-and-modify-the-risk-of-different-types-of-cancer/). All bona fide researchers can apply to use the UK Biobank resource for health related research that is in the public interest.

## ACKNOWLEDGEMENTS

We would like to thank the UKB staff and the UKB participants.

## FUNDING

The work was supported by a Brunel Research Initiative and Enterprise Fund. HAA is the recipient of a PhD studentship from the College of Health and Life Sciences, Brunel University London.

## DISCLOSURES

The authors have no conflicts of interest to declare.

## Notes

### Competing Interest Statement

The authors have declared no competing interest.

